# The need for speed: ultra-rapid high-resolution outbreak analysis in a front-line hospital microbiology laboratory

**DOI:** 10.1101/2025.02.04.25321496

**Authors:** Max Bloomfield, Sarah Bakker, Megan Burton, Kristin Dyet, Alexandra Eustace, Samantha Hutton, Jamaal Jeram, Donia Macartney-Coxson, David J. Winter, Rhys T. White

**Author notes:** Corresponding author: Rhys T. White, Institute of Environmental Science and Research, Wellington, New Zealand.

## Abstract

Many hospital laboratories have technical capacity to perform whole-genome sequencing but lack bioinformatic expertise to analyse sequence data. Sending isolates to reference laboratories creates delays that can be highly detrimental to outbreak responses. The Wellington Regional Hospital laboratory, which lacks on-site bioinformaticians, implemented real-time nanopore-based genomic surveillance that has detected several hospital outbreaks at an early stage. This has required off-site analysis, often taking weeks. Solu Genomics, a cloud-based automated bioinformatic platform, requires no bioinformatic or command-line expertise and accepts basecalled sequence files or genome assemblies. This study aimed to use Solu to replicate the analysis of two prior neonatal unit outbreaks detected by on-site genomic surveillance, as if they had occurred now, and compare the output to ‘manual’ bioinformatic analysis. Surveillance isolates that had been sequenced up until the beginning of each outbreak were loaded into Solu to replicate the background genomic data available when each outbreak occurred. The 13 methicillin-resistant Staphylococcus aureus (MRSA) and seven Klebsiella variicola outbreak isolates were then uploaded. Including upload time, each Solu analysis was completed in under 40 minutes. The phylogenetic trees generated showed distinct clustering of outbreak isolates, with overall tree topologies similar to the manual analyses. Median pairwise single-nucleotide variant distances were 12 (range 4-27) and 6 (range 1-11) for the MRSA and K. variicola outbreaks, respectively, versus 6 (range 0-14) and 18 (range 0-54) for the manual analyses. This study demonstrates that Solu Genomics can provide high-resolution, actionable outbreak analysis within minutes, without the need for bioinformatic expertise.

**Importance:** Outbreaks in healthcare settings present serious risks to patient safety and can disrupt healthcare delivery. Timely and precise detection of these outbreaks is essential for effective infection prevention and control as well as reducing patient harm and service disruptions. This study emphasises the value of an intuitive cloud-based bioinformatics platform, in overcoming a key obstacle to the widespread adoption of whole-genome sequencing in hospital laboratories: bioinformatic analyses. Our research highlights the practicality of real-time genomic monitoring in local laboratories by showing that a non-specialist platform can produce results that complement and align with manual analyses while offering significant time savings. This approach can accelerate the detection and subsequent control of outbreaks caused by organisms like methicillin-resistant Staphylococcus aureus and Klebsiella variicola, ultimately improving patient outcomes and healthcare efficiency.

## Introduction

Hospital outbreaks cause considerable patient morbidity and mortality, and huge disruption to healthcare delivery globally (1). A key determinant of an outbreak’s impact is time to detection, which directly influences the timeliness of the infection prevention and control (IPC) response (2). Outbreaks detected and accurately defined early can often be controlled with relatively little patient impact and service disruption (3, 4). Whole-genome sequencing (WGS) is a cornerstone technology for accurately detecting and controlling hospital-associated outbreaks, enabling inclusion and exclusion of organisms from clusters. Traditionally, the complexities and costs of WGS have limited its use to larger reference or research laboratories, meaning isolates require shipping off-site from hospital laboratories, imposing delays on the return of actionable results to IPC teams. However, the WGS landscape is rapidly changing. The ‘wet-lab’ aspects of WGS are increasingly accessible to front-line laboratories, with platforms such as the Oxford Nanopore Technologies (ONT) MinION™ offering the ability to perform sequencing at relatively low cost and lab space requirements, and within existing molecular scientist expertise. The ‘dry-lab’ (bioinformatic) aspects of WGS, however, remain challenging, and many laboratories lack this expertise (5). This may prevent the implementation of WGS locally, or require outsourcing of bioinformatics’ expertise, both of which cause delays to outbreak analysis and management.

To support local IPC, our laboratory introduced real-time ONT-based genomic surveillance of hospital-associated bacteria in 2022 (6). When target organisms are isolated in the laboratory, e.g., methicillin-resistant Staphylococcus aureus (MRSA) from the neonatal intensive care unit (NICU), they are added to a weekly sequencing run. Because our laboratory lacks bioinformatic expertise we employ a two-tier analysis model, whereby basic in-house analysis consists of multi-locus sequence type (MLST) determination, with transfer of sequence data to a reference laboratory for high-resolution analysis if MLST results raise suspicion of transmission events (6). This model has proven successful in detecting outbreaks (3, 4, 7), however, there are inherent delays associated with data transfer and manual analysis, meaning high-resolution analysis results may take days or weeks to return (3, 4, 7).

Recently, a secure, cloud-based, and automated bioinformatic platform called Solu Genomics (https://www.solugenomics.com/, accessed 16 December 2024) has become available, which has a range of outputs including phylogenetic analysis (8). This is operated via a Graphical User Interface (GUI) and requires no bioinformatics expertise; basecalled sequence files (or assembled genomes) are uploaded, and analysis runs automatically. This study aimed to replicate the bioinformatic analysis of two previously described outbreaks at our institution (3, 4), that were detected using proactive genome monitoring, as if they had occurred today and the laboratory had access to the Solu Genomics platform.

## Materials and Methods

### Setting

Awanui Labs Wellington (Wellington, New Zealand) is a medium-sized laboratory based on the Wellington Regional Hospital (WRH) campus. WRH serves as the tertiary referral centre for the lower North Island of New Zealand (known as Aotearoa in the Māori language). The microbiology and molecular departments of Awanui Labs Wellington (an ISO15189 accredited medical laboratory) process approximately 300,000 samples yearly. The molecular department performs a range of commercial and in-house tests, including Sanger sequencing. Since early 2022, a weekly sequencing run on the MinION™ device was performed, for the purposes described above. The initial bioinformatic analysis performed in the hospital laboratory was MLST determination using Krocus v1.0.3 (9) with default settings to query the raw fastq files (outputted from the basecalling) against either the Klebsiella PasteurMLST sequence definition database (10), or the S. aureus typing database (11) hosted on BIGSdb v1.47.0 (12). No further strain typing is performed locally, due to the lack of bioinformatics expertise.

### Previous outbreaks

Outbreak 1: in mid-2023 an outbreak of MRSA sequence type (ST)97 occurred on the WRH NICU. This was promptly detected by the genomic surveillance program (when there were only two known cases) and involved nine infants (3). In-house MLST results were available immediately, with high-resolution analysis following five weeks later. Of note, based on MLST results the outbreak was thought to involve ten infants, but core-genome single-nucleotide variant (SNV)-based analysis at the reference laboratory revealed one ST97 genome did not cluster with the outbreak strains. During the outbreak, three non-ST97 MRSA isolates were also detected. Original ONT sequence data from these 10 ST97 and three non-ST97 isolates were used for this analysis (Supplementary Materials, Table S1).

Outbreak 2: in mid-2024 an outbreak of Klebsiella variicola ST6385 occurred on the WRH NICU. This outbreak was also detected rapidly (within 48-hours of notification of an increased incidence of Klebsiella spp. bacteraemia) and involved four infants and two environmental sources (4). High-resolution phylogenetic analysis was reported four weeks later, demonstrating clustering of outbreak isolates. Original sequence data from these six isolates, plus a K. variicola ST3938 isolate detected during the outbreak were used in this analysis (Supplementary Materials, Table S2).

### Background surveillance isolates

As part of the genomic surveillance program, all MRSA isolated from the NICU from January 2022 until June 2023 (time of Outbreak 1), and all Klebsiella pneumoniae from WRH inpatients from January 2022 until April 2023 had been sequenced. Many K. pneumoniae isolates had been misidentified by conventional laboratory methods and were actually K. variicola, which is a recognised phenomenon (4). Basecalled fastq files for these NICU MRSA (n=7) and K. variicola (n=43) isolates were loaded into the Solu Genomics platform to replicate the local genomic data available when each outbreak investigation began (Supplementary Materials, Table S3).

### Sequencing and retrospective basecalling

Details on DNA extraction have been published previously (4). For isolates sequenced before March 2023, library preparation used 50nng of genomic DNA with the ONT rapid barcoding kit 96 (SQK-RBK110-96) as per the manufacturer’s instructions. The library was loaded onto a R9.4 flow cell (FLO-MIN106) and run on a MinION™ device for 20–40nhours with MinKNOW v22.10.10. After March 2023, including during the outbreaks, libraries were created using 50–100nng of genomic DNA (prepared with the rapid barcoding kit 96 (SQK-RBK114-96)), and sequenced on an R10.4.1 flow cell (FLO-MIN114) with MinKNOW v23.04.5.

Original Fast5 sequence files were converted to Pod5 using pod5 v0.3.2 (13). To replicate sequence data that would be produced had these isolates been sequenced now, Pod5 files were basecalled using Dorado v0.6.3 (14), which is equivalent to the Dorado basecall server v7.4.14 available in the standard MinKNOW install at the time of writing (14). Basecalling was carried out using the ‘super accuracy’ models, with a batch size of 2008 and a chunk size of 1000, while parameters were kept at default settings. The ‘dna_r9.4.1_e8_sup@v3.6’ model was used for the sequence data representing (i) 41 K. variicola cases collected between January 2022 and March 2023; and (ii) seven S. aureus cases collected between February-December 2022 (Supplementary Materials, Table S3). The ‘dna_r10.4.1_e8.2_400bps_sup@v4.3.0’ model was used for sequence data representing (i) seven K. variicola cases (from the outbreak April-June 2024) (Supplementary Materials, Table S2); and (ii) 13 S. aureus cases (from the outbreak June-July 2023) (Supplementary Materials, Table S1). All basecalling used a NVIDIA A100 graphics processing unit (GPU).

### Analyses using manual bioinformatic workflow

Figure 1A provides an overview of the workflow for our manual bioinformatic analyses, with further details in the Supplementary Materials. Parameters were consistent with our previous works (3).

**Figure 1.**
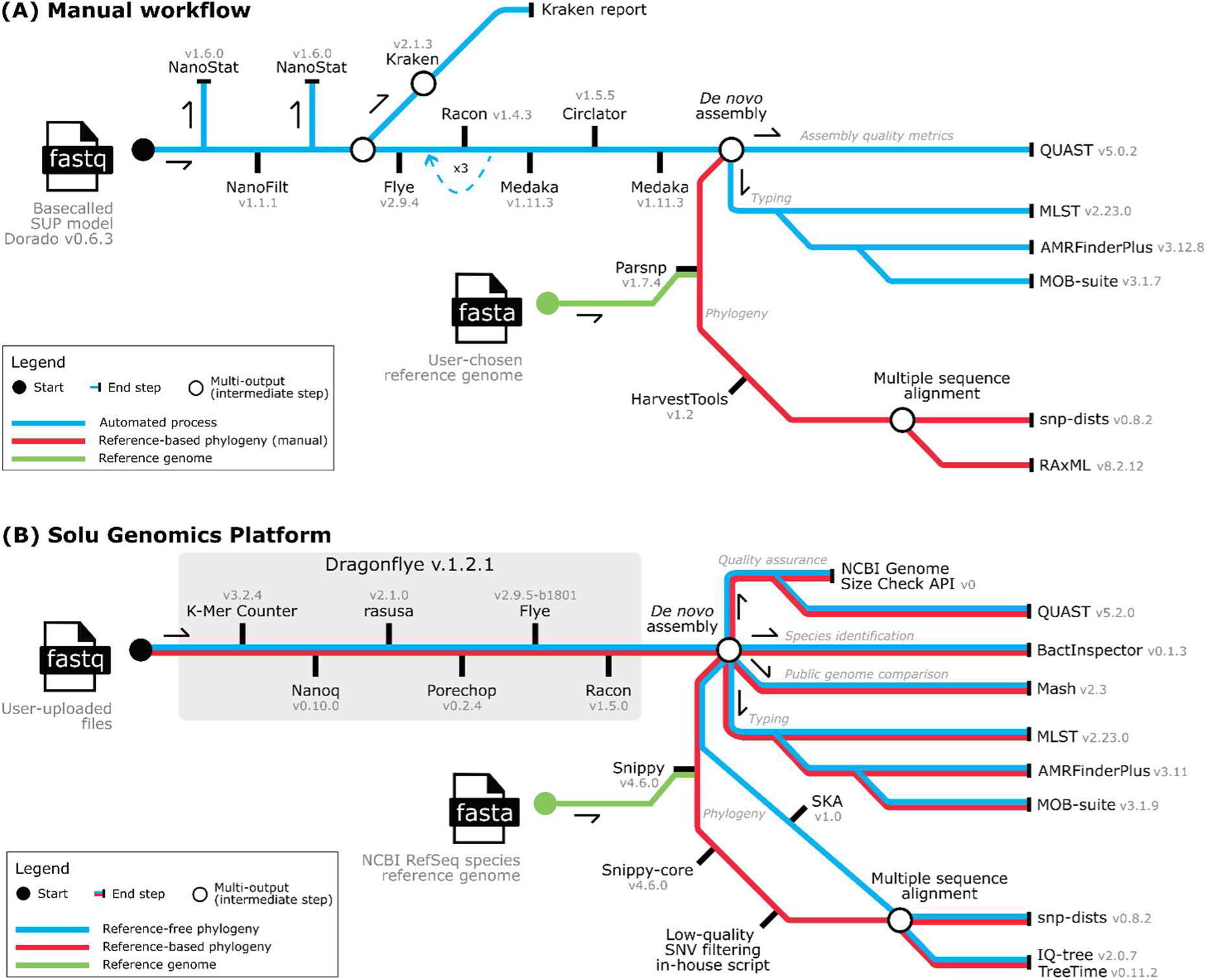
Example analysis workflow for bioinformatic analyses. (A) This ‘manual’ workflow outlines the step-by-step process for analysing basecalled sequence data files (fastq). The de novo assemblies and outputs (sequence data and assembly quality metrics, and typing data) are generated from an automated pipeline. The reference-based phylogenetic analyses is a manual step, requiring a user chosen reference genome. Pathways are color-coded to represent automated processes (blue), reference-based phylogeny (red), and reference genome integration (green). (B) This cloud-based Solu Genomics platform workflow demonstrates the step-by-step bioinformatics pipeline for processing user-uploaded basecalled sequence data files (fastq). The workflow includes both reference-free and reference-based phylogenomic analyses. The workflow produces intermediate and final outputs, including sequence alignments, phylogenies, and typing data, with color-coded pathways distinguishing reference-based phylogeny (red), reference-free phylogeny (blue), and reference data integration (green).

### Solu Genomics analysis

Fastq files from the 13 MRSA and seven K. variicola outbreak isolates were uploaded into Solu Genomics via the online GUI (https://www.solugenomics.com/, accessed 12 and 11 December 2024, respectively). All isolates from each outbreak were uploaded in bulk (sequentially), with each outbreak analysed at different times. Timestamps were taken for each isolate when the upload began, when the analysis began, and when the analysis was complete. A representation of the Solu Genomics workflow is shown in Figure 1B, with further details published (8). The reported measures of interest were: time taken to produce results; how well the results matched manual analysis; and whether the results were actionable from an IPC perspective.

### Comparing phylogenies from each workflow

The phylogenetic trees were visualised using FigTree v1.4.4 (http://tree.bio.ed.ac.uk/software/figtree/, accessed 05 November 2024). To assess congruence between the phylogenetic outputs from each workflow, for the same strains, Phylo.IO (15) was used to compare the maximum likelihood trees.

### High-resolution read-mapping approach to variant calling and SNV density analysis

To compare the performance of the Solu Genomics platform using sequence data basecalled with Dorado v0.6.3 (corresponding to the Dorado integrated with MinKNOW v24.06.16) to the manual analysis using the most up-to-date standalone Dorado, Pod5 files were basecalled using Dorado v0.8.2. Basecalling (using a NVIDIA A100 GPU) was carried out using the ‘super accuracy’ model dna_r10.4.1_e8.2_400bps_sup@v5.0.0, with a batch size of 2008 and a chunk size of 1000, while parameters were kept at their default settings.

We used the complete chromosomal genome sequences of S. aureus strain sa230627barcode06 (GenBank: CP143800), representing the ST97 outbreak, and K. variicola strain kv240612_barcode19 (GenBank: CP165787), representing the ST6385 outbreak, as references to call SNVs. The quality-trimmed and filtered reads from the five other K. variicola ST6385 isolates and eight other S. aureus ST97 isolates were aligned to the respective reference genomes (kv240612_barcode19 or sa230627barcode06) using minimap2 v2.28 (16, 17) with the ‘map-ont’ preset for long-read data. The resulting Sequence Alignment/Map (SAM) file was converted to Binary Alignment/Map (BAM) format using SAMtools v1.20 (18). The BAM file was sorted and indexed to facilitate downstream analysis. Variant calling was conducted using Clair3 v1.0.8 (19) with the ‘r1041_e82_400bps_sup_v500’ model, and flags ‘--include_all_ctgs’, ‘--no_phasing_for_fa’, and ‘--haploid_precise’. Variants were filtered using BCFtools v1.20 (20) to exclude those with a quality score below 30. Consensus sequences for each genome were created by integrating strain-specific SNVs into the respective reference chromosomes (kv240612_barcode19 or sa230627barcode06). SNVs were extracted from the concatenated pseudo-genome alignment using SNP-sites v2.4.1 (21), generating a variant-only alignment for phylogenetic analyses. RAxML v8.2.12 (22) built phylogenetic trees using the maximum-likelihood method with GTR-GAMMA correction (optimising 20 distinct, randomised maximum-parsimony trees). The calculation of pairwise SNV distances were executed using snp-dists v0.6.3 (23).

## Results

The isolates and corresponding quality control metrics for each outbreak are shown in Table 1. Sequencing data for the isolates had sufficient estimated depth (18× to 384×) and median read lengths (783 bp and 4,709 bp) for genomic analyses. Overall, the median Phred Quality Score across all samples was Q18.7 (interquartile range: Q17.2 to Q19.7; range Q15.8 to Q20.6), indicating an accuracy >98.6%, with an estimated 1 in ∼76 base calls likely to be incorrect.

**Table 1.**
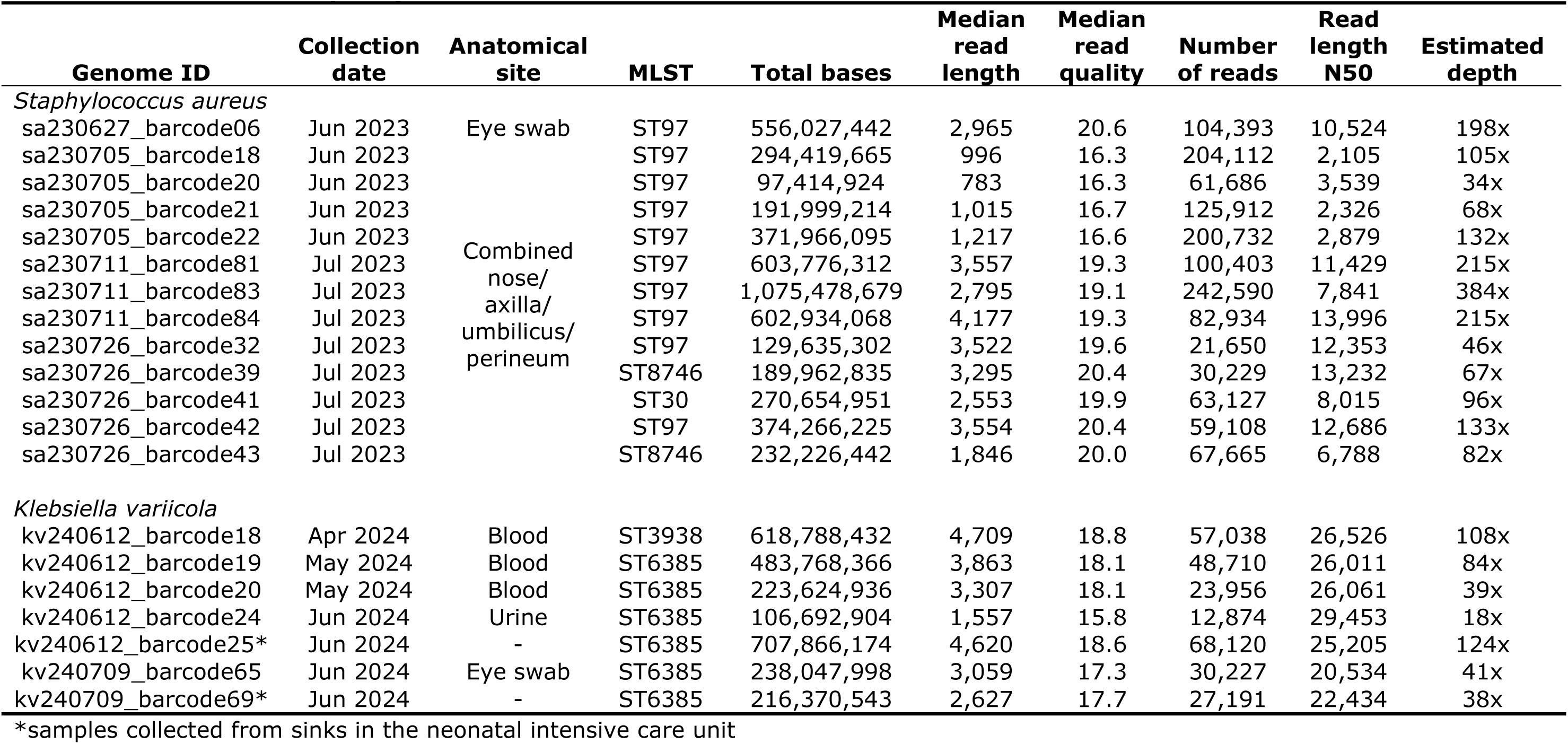
General isolate/quality control metrics for each outbreak.

### Replicating and analysing the MRSA outbreak

The Solu Genomics platform completed the analysis in 27 minutes (6 minutes upload and 21 minutes analysis; Supplementary Materials Figure S1), using a reference-based approach (Figure 2) with strain NCTC 8325 (GenBank: CP000253) as a reference to call SNVs.

**Figure 2.**
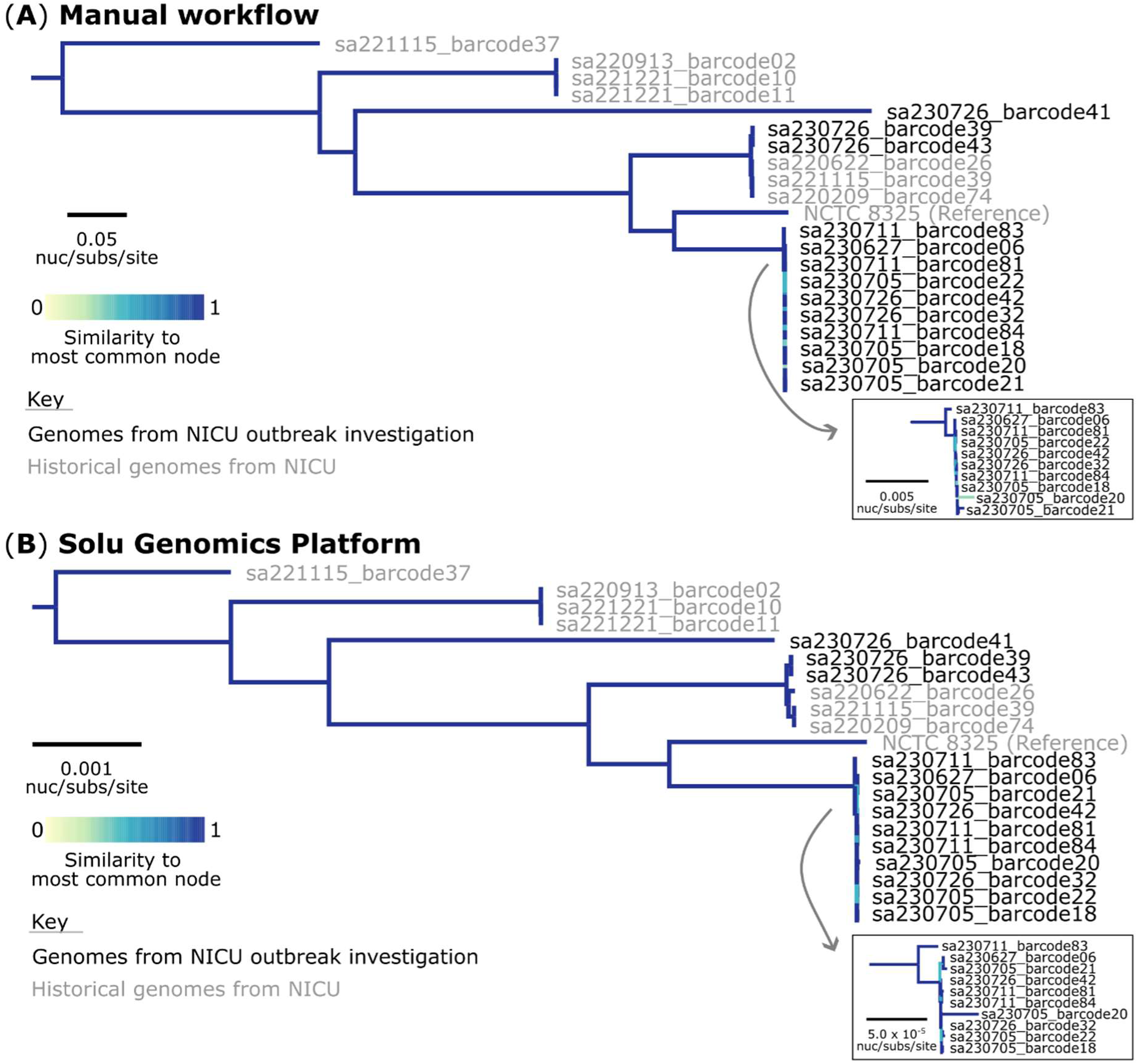
Comparison of sequencing analysis workflows for the Staphylococcus aureus outbreak. (A) Maximum likelihood generated from the manual bioinformatic analyses. The phylogeny was inferred from 51,942 core-genome single-nucleotide variants (SNVs) from 21 assembled genomes. SNVs were derived from a core-genome alignment of 1,632,539 bp. In both phylogenetic trees, SNVs were called against the 2,821,361 bp chromosome of strain NCTC 8325 (GenBank: CP000253). (B) Maximum likelihood generated from the Solu Genomics cloud-based platform. The phylogeny was inferred from 48,855 core-genome SNVs from 21 assembled genomes. The multiple sequence alignment was 2,821,361 bp in length. The trees are rooted to the actual root by S. aureus sequence type (ST)93 (sample: sa221115_barcode37; SRA: SRR31828390). The colour of the branches represents the comparison metric. A score of 1 denotes that the subtree structure of the node is identical to the subtree structure of its best corresponding node.

Side-by-side tree comparison and degree of correspondence between the nodes using Jaccard scores (Figure 2) showed that both approaches produced highly concordant phylogenetic trees with nearly identical topological structures. Minor variations in branch colouring within the ST97 clade reflected subtle differences in similarity scores to the most common node, but overall tree topology and clustering patterns were consistent. The sporadic ST97 isolate detected during the outbreak formed an outgroup in both analyses (isolate sa230711_barcode83, Figure 2 inset).

When examining the ST97 outbreak isolates more closely (Supplementary Materials Figure S2), we observed differences in the estimated genetic diversity between the two approaches. The manual workflow using updated Dorado models and a closely matched reference genome found a median of 6 SNVs (IQR: 5 to 10; range: 0 to 14; Supplementary Materials Figure S2A), while the Solu Genomics workflow identified a median pairwise distance of 12 SNVs between isolates (IQR: 8 to 19; range: 4 to 27; Supplementary Materials Figure S2B).

### Replicating and analysing the K. variicola outbreak

The Solu Genomics platform completed this analysis in 32 minutes (four minutes upload, 28 minutes analysis; Supplementary Materials Figure S3). Unlike the S. aureus analysis, the Solu Genomics platform lacked a K. variicola reference genome (at the time of analysis). Consequently, a Split K-mer Analysis (SKA) approach was used (Figure 1B) to identify genetic variations associated with the outbreak. Our manual reference-based workflow (Figure 3A) and the SKA-based Solu Genomics analysis (Figure 3B) produced similar tree topologies, particularly in the clustering of the NICU outbreak investigation genomes relative to historical isolates. Phylogenetic relationships between isolates were preserved across both analyses, with the outbreak isolates forming a distinct cluster. When examining the ST6385 outbreak isolates more closely (Supplementary Materials, Figure S4), we observed differences in the estimated genetic diversity between the SKA (reference-free) and reference-based approaches. The manual workflow found a median of 18 SNVs (IQR: 2 to 45; range: 0 to 48; Supplementary Materials, Figure S4A), while the Solu Genomics workflow identified a median SKA distance of 6 SNVs between isolates (IQR: 5 to 7; range: 1 to 11; Supplementary Materials, Figure S4B).

**Figure 3.**
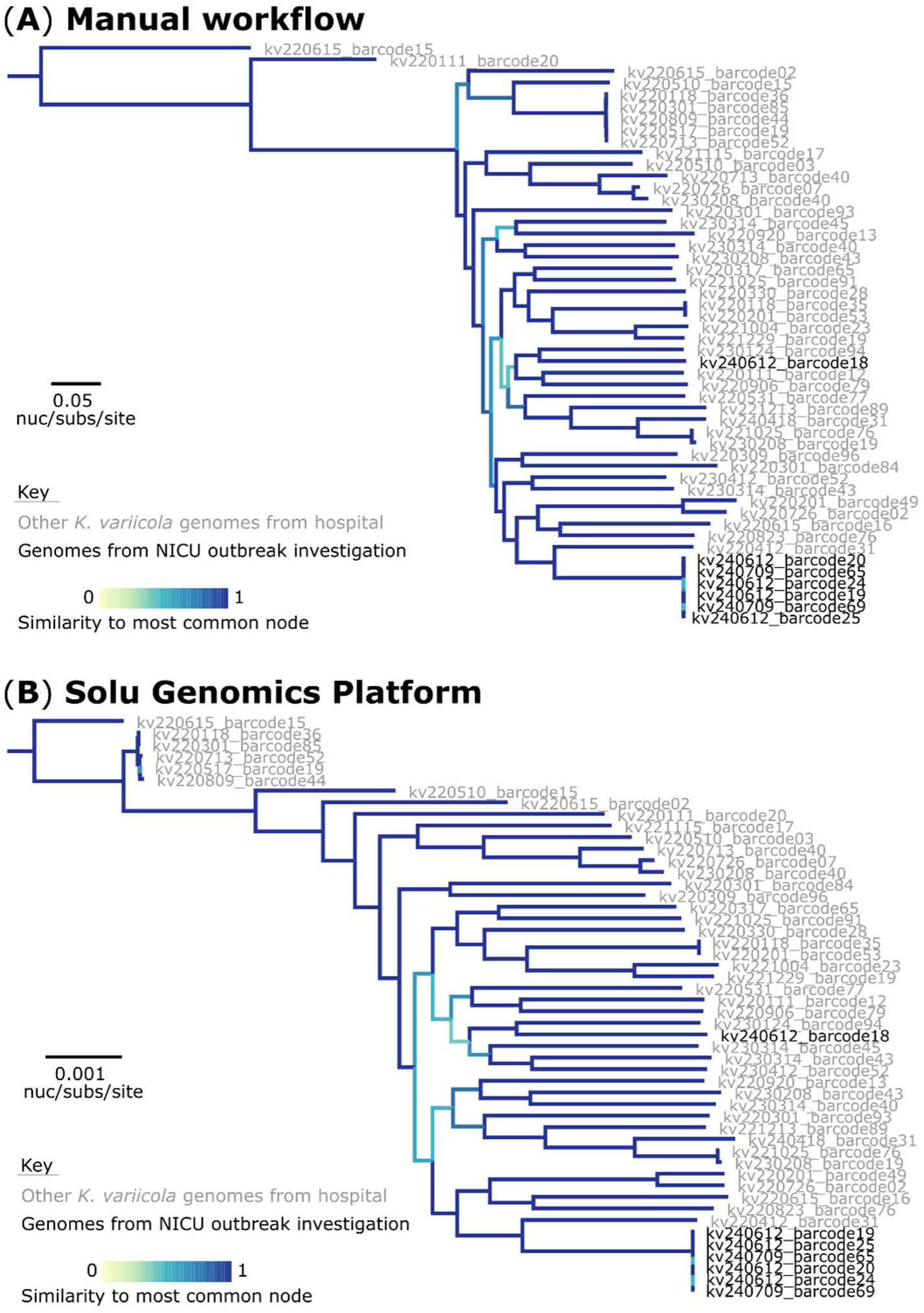
Comparison of sequencing analysis workflows for the Klebsiella variicola outbreak. (A) Maximum likelihood generated from our established workflow. The phylogeny was inferred from 182,753 core-genome single-nucleotide variants (SNVs) from 50 assembled genomes. SNVs were derived from a core-genome alignment of 4,331,140 bp and are called against the 5,500,654 bp chromosome of sample kv240612_barcode19 (GenBank: CP165787). (B) Maximum likelihood generated from the Solu Genomics cloud-based platform (reference free). The phylogeny was inferred from 87,600 core-genome SNVs from 50 assembled genomes. The multiple sequence alignment was 3,327,801 bp in length. The trees are rooted to the actual root by K. variicola sequence type (ST)3081 (sample: kv220615_barcode15; SRA: SRR30085197).

## Discussion

In this reanalysis of two recent hospital outbreak datasets, high-resolution and actionable results were produced rapidly using the cloud-based Solu Genomics platform. In each outbreak, the time from the beginning of the upload until results was approximately 30 minutes, with no user input required other than the initial basecalled fastq upload procedure. The results produced would have been directly actionable from an IPC perspective had they occurred as part of a ‘live’ outbreak investigation, with outbreak isolates forming distinct clusters in the phylogenetic trees, compared to background surveillance isolates. Importantly, other isolates collected during the outbreak investigations that were unrelated formed clear outgroups in the Solu analyses.

The Solu phylogenetic tree topologies, whilst not identical, were similar to the manual analyses, particularly concerning the outbreak clades. The Solu analysis produced larger SNV distances for the MRSA outbreak which may reflect the more accurate basecalling model and closely matching reference genome used for the manual analysis (24). Conversely, the Solu analysis produced smaller SNV distances for the K. variicola outbreak, likely due to recombination events between outbreak isolates (4). These events had a greater impact on the SNV distances in the manual analysis compared to the Solu analysis. This may reflect the SKA approach used in the Solu analysis, which is designed to analyse closely-related genomes (25). Additionally, SKA appears to underestimate SNV distances when compared to core-genome SNV analysis (26). Despite the SNV differences between the two analytical approaches, the overall conclusions from an IPC perspective were the same, with the outbreak isolates forming clear clusters compared to background surveillance isolates. This highlights an advantage of real-time surveillance where potential outbreaks can be contextualised in light of local genomic epidemiology.

When paired with ONT sequencing, which allows utilisation of sequence data in real-time, this analysis model would potentially allow same-day (from the start of genomic library preparation) high-resolution outbreak investigation (6). In addition to the speed of results, the simple cloud-based GUI operation makes it accessible to front-line clinical laboratories that can perform the ‘wet-lab’ aspects of sequencing but lack bioinformatics expertise. Bringing WGS with high-resolution analysis to the hospital laboratory setting has the potential to enable extremely agile data-driven IPC responses to emerging hospital-based outbreaks, removing many of the delays associated with the transfer of samples or data off-site. In our experience, direct communication between IPC and the hospital laboratory has facilitated highly responsive outbreak analysis (3, 4), with access to rapid high-resolution analysis, as shown here, offering the potential for further improvement. Solu Genomics is well-suited to real-time genomic surveillance, as newly sequenced isolates can be added to existing phylogenies, as was done in this study.

A limitation of this study was that it involved only two small outbreak datasets, which may limit generalisation of results to other situations and sequencing platforms, and larger outbreaks. However, the Solu Genomics pipeline has been validated on other datasets, and analysis of isolates occurs in parallel on the platform, so the rate-limiting factor for analysis of larger outbreaks would likely relate to upload time; analysis time would likely be similar (8). Whilst access to automated high-resolution analysis offers significant opportunity for front-line laboratories to provide WGS services, like any bioinformatic analysis, considerable care needs to be taken when interpreting results. Laboratories unfamiliar with WGS analysis outputs may misinterpret results, which could lead to incorrect IPC actions. For example, Solu Genomics employs a single reference genome for each species, which, if not well-matched to the genomes being analysed, could in some instances create spurious results with isolates appearing more or less closely related than they truly are (24). We believe that personnel familiar with WGS outputs should be available to interpret results, along with bioinformatics support to provide further analysis where necessary. In our context, we see Solu as having considerable potential for empowering hospital laboratories and IPC teams to rapidly respond to potential outbreaks, but believe that oversight from a centralised bioinformatic facility will be critical. The platform is well-suited to this, with the ability for multiple laboratories to upload data and share results, and could support a decentralised sequencing approach, whereby multiple ‘spoke’ laboratories produce and upload sequence data to the platform, with a central ‘hub’ facility providing coordinated oversight of results (27). Challenges for implementation will include quality assurance of the ‘wet-lab’ aspects, local validation of the overall process, and jurisdictional privacy and data sovereignty considerations.

The main strength of this study was that we were able to replicate real-world conditions, whereby sequence data were generated using current, readily-available sequencing technology in a hospital laboratory setting. We basecalled prior sequence data using the Dorado model that is equivalent to what is available in the current standard MinKNOW install (at the time of writing), so other laboratories that are performing ONT sequencing using standard parameters could reasonably expect to produce similar results to what are presented here.

In summary, this study has demonstrated the ability of the Solu Genomics platform to provide ultra-rapid high-resolution analysis of hospital-based outbreaks, with sequence data that had been generated in a front-line hospital laboratory. This represents a significant advance towards wider access to local, democratised WGS analysis, offering a combination of speed and precision currently unavailable to most hospital laboratories and IPC teams. Rapid, data-driven responses to potential outbreaks offers an opportunity to minimise their impact on patients and healthcare delivery.

## Author statements

## Authors and contributions

Conceptualisation: MBl and RTW; investigation: MBl and RTW; funding acquisition: DMC, DJW, and MBl; formal analysis: RTW; wet-lab experiments: SB, MBu, AE, SH, JJ; data analysis: RTW; data curation: MBl and RTW; writing (original draft preparation): MBl and RTW; writing (review and editing): MBl, SB, Mbu, KD, AE, SH, JJ, DMC, DJW, and RTW. All authors have read and agreed to the published version of the manuscript.

## Ethics statement

The analysis and reporting of this outbreak constituted an ‘audit or related activity’ as per New Zealand Health and Disability Ethics Committees, so did not require review. Ethics waiver was granted under New Zealand Health and Disability Ethics Committee requirements.

## Data Availability

The Oxford Nanopore Technologies raw sequence read data, basecalled using Dorado v0.6.3, have been deposited in the National Center for Biotechnology Information (NCBI) sequence read archive (SRA, https://www.ncbi.nlm.nih.gov/sra). The sequence data for Staphylococcus aureus are available under BioProject accession numbers PRJNA1046639 (SRA: SRR31828393 to SRR31828405), PRJNA1090129 (SRA: SRR31828386 to SRR31828392), and PRJNA1144171 (SRA: SRR31828385). The Klebsiella variicola sequences are available under BioProject accession numbers PRJNA1142676 (SRA: SRR31834905 to SRR31834938), PRJNA1142678 (SRA: SRR31834945 to SRR31834952), and PRJNA1142680 (SRA: SRR31834953 to SRR31834960).

## Acknowledgements

We acknowledge the facilities, and the scientific and technical assistance of staff at Awanui Labs, formerly known as Southern Community Laboratories (Wellington, New Zealand). We also acknowledge the Antibiotic Reference Laboratories at the Institute of Environmental Science and Research (ESR, Porirua, New Zealand). We thank the following staff at ESR for their valuable feedback: Zuyu Yang (Kenepuru Science Centre) and Ernest Williams (Wallaceville Science Centre). This research was made possible by the Computational Science Team and the High-Performance Compute (HPC) platform at ESR, with specific thanks to Russell Smithies and Shane Sturrock for HPC. We would also like to thank the Solu team for access to the Solu Genomics platform.

## Data Availability Statement

The Oxford Nanopore Technologies raw sequence read data, basecalled using Dorado v0.6.3, have been deposited in the National Center for Biotechnology Information (NCBI) sequence read archive (SRA, https://www.ncbi.nlm.nih.gov/sra). The sequence data for Staphylococcus aureus are available under BioProject accession numbers PRJNA1046639 (SRA: SRR31828393 to SRR31828405), PRJNA1090129 (SRA: SRR31828386 to SRR31828392), and PRJNA1144171 (SRA: SRR31828385). The Klebsiella variicola sequences are available under BioProject accession numbers PRJNA1142676 (SRA: SRR31834905 to SRR31834938), PRJNA1142678 (SRA: SRR31834945 to SRR31834952), and PRJNA1142680 (SRA: SRR31834953 to SRR31834960). The authors confirm that all supporting data protocols have been provided in the article or supplementary data files.

