## Supplementary Materials for "The need for speed: ultra-rapid high-resolution outbreak analysis in a front-line hospital microbiology laboratory"

**Affiliations(s)**

[1]Awanui Labs Wellington, Department of Microbiology and Molecular Pathology, Wellington 6021, New Zealand

[2]Te Whatu Ora/Health New Zealand, Infection Prevention and Control, Capital, Coast & Hutt Valley, Wellington 6021, New Zealand

[3]Institute of Environmental Science and Research, Health Group, Porirua 5022, New Zealand

**Corresponding author and email address**


<u>**This file includes the following:**</u>

**Supplementary Methods**.

**Supplementary Figure S1**. Estimated sequencing depth against computational time for *Staphylococcus aureus* genomic analysis

**Supplementary Figure S2**. Comparison of phylogenetic analysis for the *Staphylococcus aureus* sequence type (ST)97 outbreak.

**Supplementary Figure S3**. Estimated sequencing depth against computational time for *Klebsiella variicola* genomic analysis.

**Supplementary Figure S4**. Comparison of phylogenetic analysis for the *Klebsiella variicola* sequence type (ST)6385 outbreak.

### Supplementary Methods

*Analyses using established bioinformatic workflow*

Basecalled nanopore reads underwent quality assessment (NanoStat v1.6.0 (1)) and filtering (NanoFilt v1.1.1 (1)), with 52 nucleotides trimmed from read ends and a Q7 quality threshold. Taxonomic profiling used Kraken v2.1.3 (2) with the NCBI RefSeq Standard database (https://benlangmead.github.io/aws-indexes/k2, accessed on 02 January 2025 (3)). The database contained references for archaea, bacteria, human, viruses, plasmids, and the 'UniVec core' subset of the UniVec database (a database of vector, adaptor, linker and primer sequences). *De novo* assembly was performed using Flye v2.9.4 (4, 5) (genome size: 2.8 Mb for *Staphylococcus aureus* or 5.5 Mb for *Klebsiella variicola*; three polishing iterations). Read alignment was done with minimap2 v2.28 (6, 7) (configured for long-read data with the 'map-ont' preset), followed by polishing to correct single-nucleotide variants (SNVs) and insertions and deletions (INDELs) using racon v1.4.3 (8) and Medaka v1.11.3 (9). After circularisation with Circlator v1.5.5 (10) and final Medaka polishing, assembly quality was assessed using QUAST v5.0.2 (11). Detailed parameters are consistent with our previous work (12). Strain typing included multi-locus sequence typing (MLST) analysis using MLST v2.23 (13) with default settings to query the assemblies against either the *Klebsiella* PasteurMLST sequence definition database (14), or the *S. aureus* typing database (15) hosted on BIGSdb v1.47.0 (16). Virulence genes, acquired antibiotic resistance genes, and mutations conferring resistance to antibiotics were identified using AMRfinderplus v3.12.8 with database version 2024-01-31.1 (17). MOB-suite v3.1.7 was used to identify plasmid replicons (18). Genome assemblies were aligned to create a core-genome alignment using Parsnp v1.7.4 (19). Resulting SNV alignments were used to reconstruct phylogenies. RaxML v8.2.12 (20) built phylogenetic trees using the maximum-likelihood method with GTR-GAMMA correction (optimising 20 distinct, randomised maximum-parsimony trees before adding 1,000 bootstrap replicates).

**Supplementary Figures**

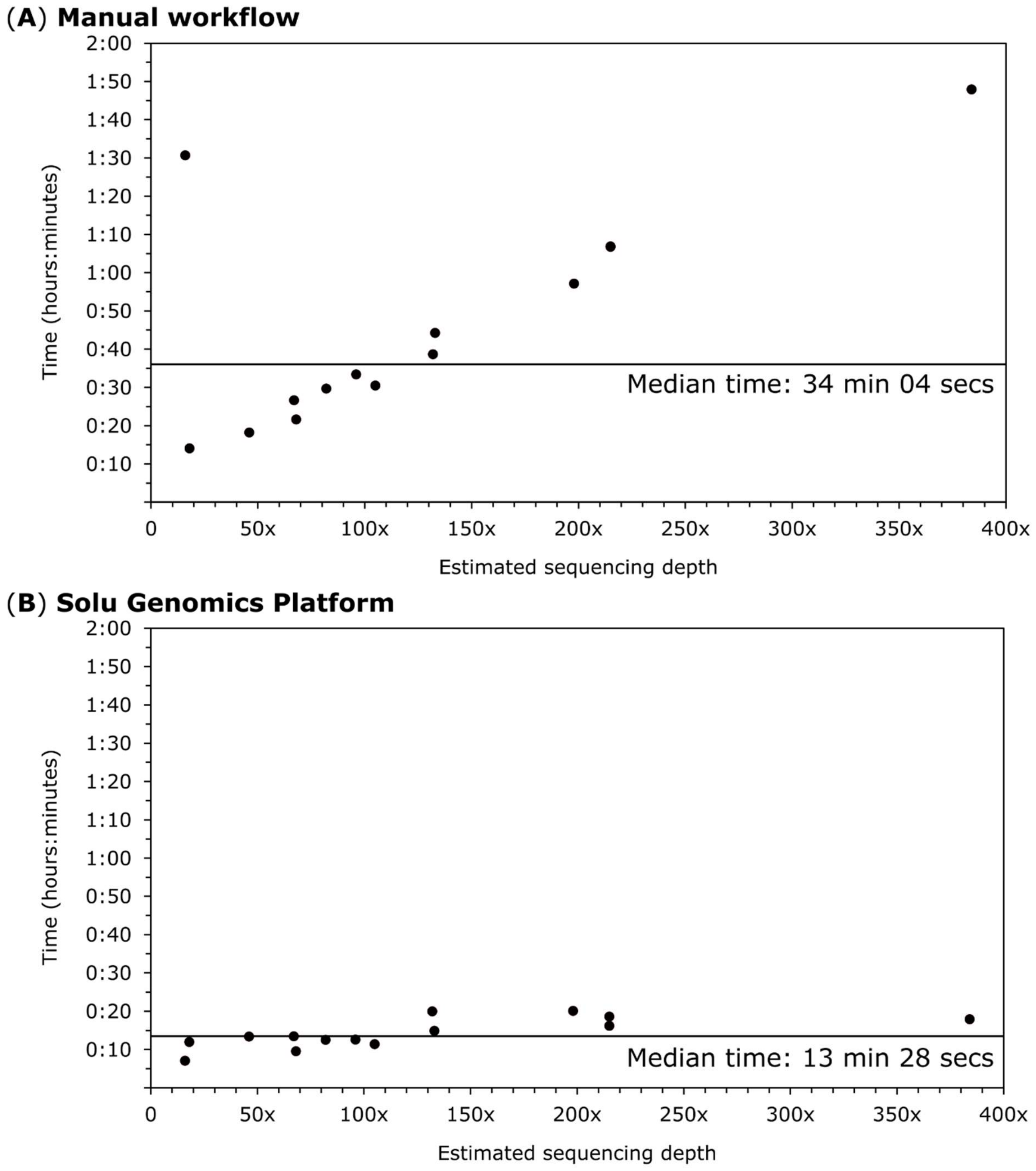


**Supplementary Figure S1. Estimated sequencing depth against computational time for *Staphylococcus aureus* genomic analysis.** Processing times for the (**A**) manual bioinformatic analyses, and (**B**) Solu Genomics cloud-based platform. Horizontal lines indicate median times for each process.

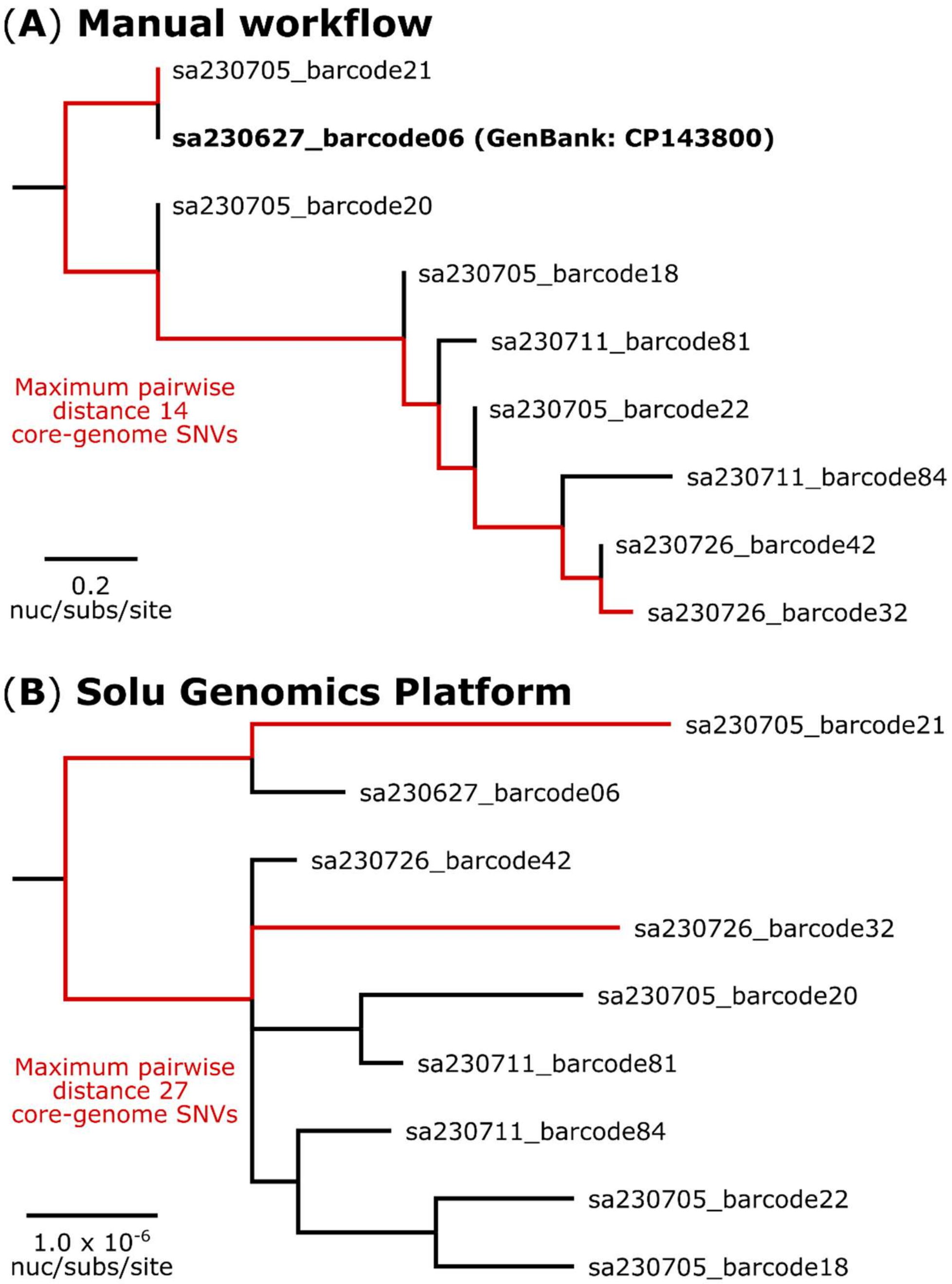


**Supplementary Figure S2. Comparison of phylogenetic analysis for the *Staphylococcus aureus* sequence type (ST)97 outbreak**. (**A**) Maximum likelihood generated from the manual bioinformatic analyses. The phylogeny was inferred from 16 core-genome single-nucleotide variants (SNVs) from nine genomes. The multiple sequence alignment was 2,753,159 bp in length. SNVs were called against the 2,753,159 bp chromosome of strain sa230627barcode06 (GenBank: CP143800). (**B**) Maximum likelihood generated from the Solu Genomics cloud-based platform. The phylogeny was inferred from 48,855 core-genome SNVs from 21 assembled genomes. The multiple sequence alignment was 2,821,361 bp in length. SNVs were called against the 2,821,361 bp chromosome of strain NCTC 8325 (GenBank: CP000253). While the species-level tree is not shown, this zoomed-in view highlights only the ST97 outbreak genomes.

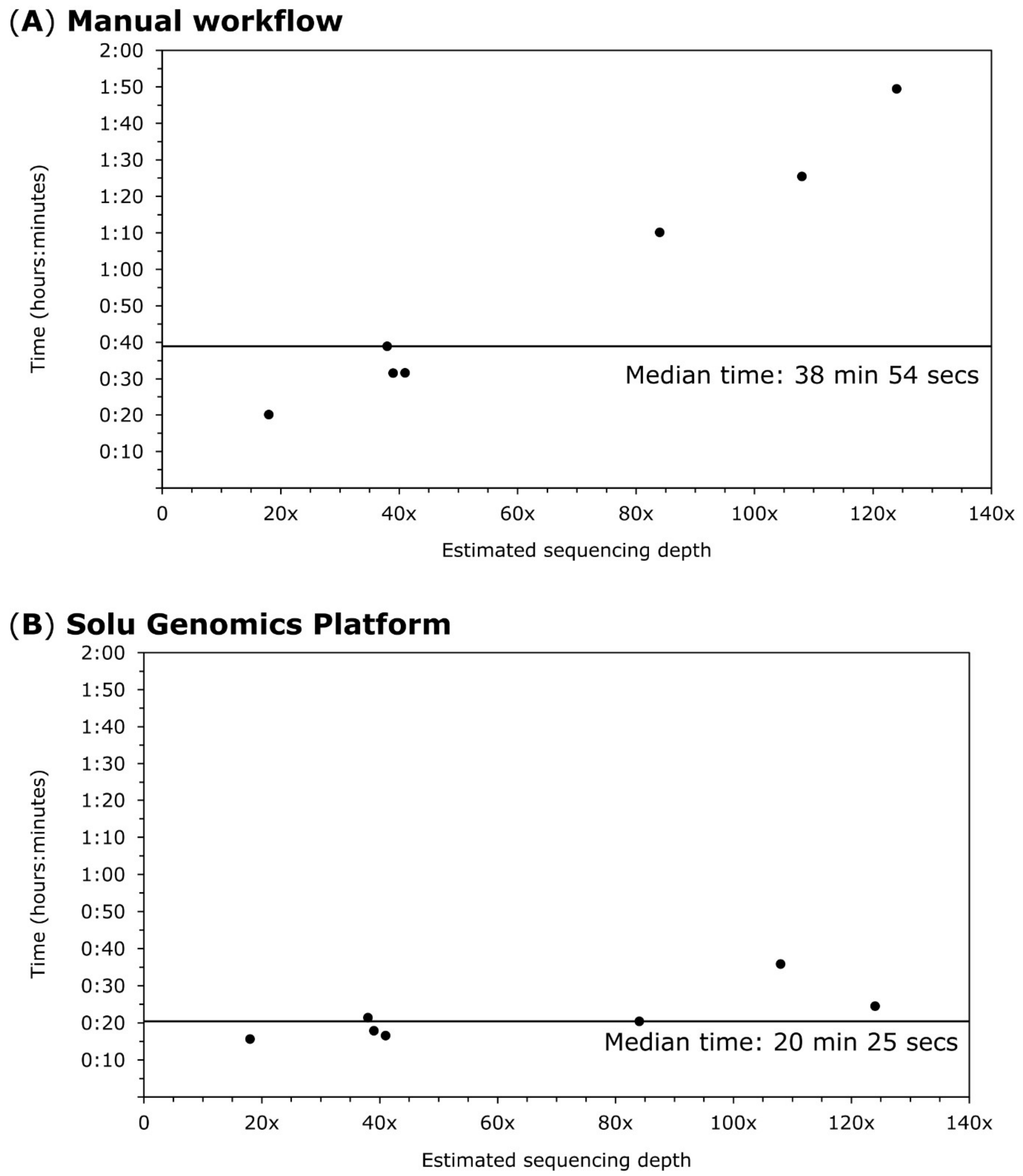


**Supplementary Figure S3. Estimated sequencing depth against computational time for *Klebsiella variicola* genomic analysis.** Processing times for the (**A**) manual bioinformatic analyses, and (**B**) Solu Genomics cloud-based platform. Horizontal lines indicate median times for each process.

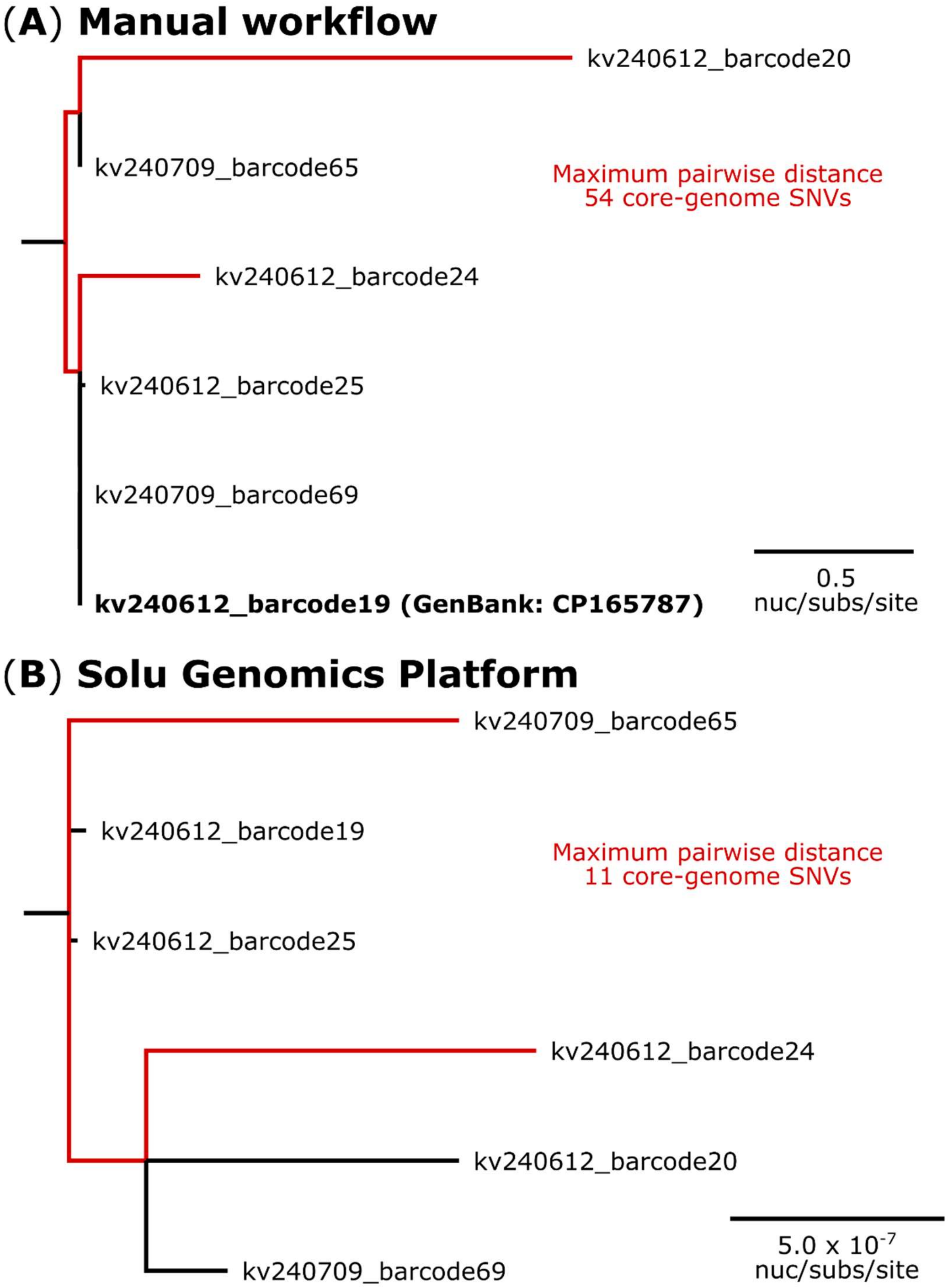


**Supplementary Figure S4. Comparison of phylogenetic analysis for the *Klebsiella variicola* sequence type (ST)6385 outbreak**. (**A**) Maximum likelihood generated from the manual bioinformatic analyses. The phylogeny was inferred from 57 core-genome single-nucleotide variants (SNVs) from six genomes. The multiple sequence alignment was 5,500,654 bp in length. SNVs were called against the 5,500,654 bp chromosome of strain kv240612_barcode19 (GenBank: CP165787). (**B**) Maximum likelihood generated from the Solu Genomics cloud-based platform (reference free). The phylogeny was inferred from 87,600 core-genome SNVs from 50 assembled genomes. The multiple sequence alignment was 3,327,801 bp in length. While the species-level tree is not shown, this zoomed-in view highlights only the ST6385 outbreak genomes.
